# HAS COUNTRYWIDE LOCKDOWN WORKED AS A FEASIBLE MEASURE IN BENDING THE COVID-19 CURVE IN DEVELOPING COUNTRIES?

**DOI:** 10.1101/2020.06.23.20138685

**Authors:** Khondoker Nazmoon Nabi, Md. Robiul Islam

**Affiliations:** Department of Mathematics, Bangladesh University of Engineering and Technology (BUET), Dhaka-1000, Bangladesh; Department of Computer Science and Engineering, Green University of Bangladesh, Dhaka-1207, Bangladesh

**Keywords:** COVID-19, growth factor, effective reproduction number, disease generation interval, disease incidence, daily cumulative index, lockdown effectiveness

## Abstract

In the absence of any effective vaccine and clinically proven treatment, experts thought that strict lockdown measures could be an effective way to slow down the spread of novel coronavirus. Despite the strict lockdown measures in several developing countries, the number of newly infected cases is getting unbridled as time progresses. This anomaly ignites questions about the effectiveness of the prolonged strict confinement measures. In light of the above view, with an aim to find the answer to this question, trends of four epidemiological parameters: growth factor of daily reported COVID-19 cases, daily incidence proportion, daily cumulative index and effective reproduction number in five developing countries named Bangladesh, Brazil, Chile, Pakistan and South Africa have been analysed meticulously considering the different phases of their national lockdowns. Any compelling evidence has not been found in favor of countrywide lockdown effectiveness in the above-mentioned countries. Numerical results illustrate that stringent nationwide lockdown measures have failed bringing the epidemic threshold (*R*_*e*_) of COVID-19 under unity. In addition, citizens of the aforementioned countries have been struggling with catastrophic socio-economic consequences due to prolonged confinement measures. Our study suggests that a new policy should be proposed for developing countries to battle against future disease outbreaks ensuring a perfect balance between saving lives and confirming livelihoods.

## 1. Introduction

On the eve of January, 2020, Chinese scientists identified a new respiratory virus named the novel coronavirus which has a virulent nature. With an intention to rein in the spread of the virus, an unprecedented aggressive lockdown has been imposed in Wuhan and in other cities in Hubei without any delay. Though this massive scale confinement strategy was discouraged by many experts in the primitive stage, as time progresses this policy has been applauded as an effective model of lockdown Lau et al., (2020). Meanwhile, the World Health Organisation (WHO) announced a global health emergency considering the severe transmissibility of COVID-19 on January 30, 2020 Nabi (2020).

In the wake of the outbreak of COVID-19 pandemic, numerous individuals who are living abroad mainly for work or travel purpose started returning back to their native countries promptly. This uncontrollable influx of arriving passengers from COVID-19 affected countries played a significant role in spreading COVID-19 transmission into the respective local population comprehensively Guilherme et al., (2020). The scenario started to exacerbate gradually because of inadequate screening of on-arrival suspected cases. Due to under-resourced and fragile health care systems in several cases, most of the developing countries failed to launch emergency plan initially to battle against this pandemic. Owing to underreporting on the early stage of the disease outbreak and scarcity of medical equipment and diagnostic kits to identify the potential spreaders, these countries failed to halt the initial outbreak of the virus Nabi (2020).

In the absence of any effective vaccine and clinically established treatment, any affected country could think of only two possible strategies to bring a control over the pandemic at an early stage of the outbreak Gollier (2020). Firstly, the entire population could be exposed to the virus with an aim to develop herd immunity in the community in a progressive way. However, in this way a targeted population (often elderly people) could die which would be highly unethical. Another strategy could be to enforce non-pharmaceutical measures like strict lockdown measures, social distancing, mandatory self or home quarantine, wearing masks and so on. Most of the countries even developing countries are no exceptions, opted for the second option without planning much about future economic shockwaves of the pandemic. With an eye towards stanching the spread of the virus, stay-at-home orders, prohibiting large gatherings of individuals and closures of all types of educational institutions and non-essential business have been imposed comprehensively. As a time-tested strategy for battling against COVID-19, restrictions on traveling have been imposed by the respective governments in developing countries. Maximum countries issued centralized quarantine orders for fourteen days to the inhabitants who have been tested positive or have had contact history with infected individuals. Stringent mask-wearing and strict physical distancing have been promoted as mass awareness program to curtail the risk of COVID-19 transmission. With an aim to ensure smooth supply chains for essential services, most of the governments allowed transportation of essential goods. Importantly, government officials continued allowing restrictive connectivity between country’s capital city and other cities amid the coronavirus outbreak as capital city always plays a remarkable role in a developing country’s economy Kamruzzaman (2020, May 28); Rivers & Gallón (2020, June 3); Sajid & Latif (2020, March 24). As a consequence, community transmission began to show up in a large scale in the developing countries.

To date, most of the developing countries are struggling hard to cope with the unforeseen socio-economic aftermaths of harsh confinement policies compared to the developed countries. Developing countries could face a net income loss of US 220 billion according to a report of the UN Development Programme (UNDP) United Nations (2020). Daily wage workers who are considered as the most vulnerable section of developing countries, have been struggling to meet up their daily income since the beginning of the outbreak. In addition, such massive income losses could severely affect their lifestyle as they do not possess any basic food security at all. For instance, some recent studies have enlightened the fact that the agro-based industries and different service sectors in Bangladesh are incurring a loss of about US 0.338 billion on daily basis. Importantly, a loss of about US 2.51 billion had been incurred by the ready-made garments sector, which is the key driving force of the country’s entire export earnings Ahmed (2020, June 10). Moreover, small businesses and startups being the worst hit are expected to incur a loss of around US 582 billion amid this pandemic Mahmood (2020, May 30). According to another research findings, the economic fallouts from the global pandemic could increase global poverty by affecting as much as half a billion people, or 8% of the global population. As a consequence, it would be really difficult to achieve the Sustainable Development Goals (SDGs) 2030 on no poverty and zero hunger Mahmood (2020, May 30). At present, with an aim to revive country’s nearly crippled economy, governments of several developing countries have been compelled to lift lockdown restrictions amid continuous spikes in both new COVID-19 cases and deaths. This seemingly ignites a fact that something went wrong in these developing countries which requires broad-ranging research.

In light of the above view, the main objective of this study is to quantify the impact and effectiveness of widespread lockdowns in five developing countries named Bangladesh, Brazil, Chile, Pakistan and South Africa ICQI (2020) using four insightful epidemiological parameters: growth factor of daily infected cases, daily disease incidence proportion, daily cumulative index and effective reproduction number.

## 2. Materials and methods

In this study, trends of four significant epidemiological metrics: growth factor (GF), daily incidence proportion (DIP), daily cumulative index (DCI) and effective reproduction number have been analysed to take a deeper look into the transmission dynamics of COVID-19 in five developing countries in the world: Bangladesh, Brazil, Chile, Pakistan, and South Africa. Daily updates of confirmed COVID-19 cases for the abovementioned countries have been collected from a trusted online repository Our World in Data COVID-19 Dataset (2020). The main reason for selecting the above-mentioned developing countries is governments of these countries have been trying earnestly to slow down the spread of the novel coronavirus by imposing strict lockdown measures from the early stage of the outbreak. However, these countries have been witnessing new records of infections and deaths with the passage of time in spite of imposing stringent confinement measures by sacrificing all kinds of economic activities. Henceforth, the effectiveness of countrywide lockdown in developing countries for a prolonged period has become a burning question for research communities as well as policymakers Meunier (2020).

Growth factor (GF) of the daily infected cases of COVID-19 has been used in this study as an insightful metric to understand the progression of a outbreak in any country Tang & Wang, (2020). A 5-day moving average of the daily infected cases of COVID-19 has been applied to calculate the growth factor of daily infected cases. This can be estimated by taking the average of the values of the observing day data, the previous two days data and the future two days data. It has been found that the number daily reported COVID-19 cases is somewhat noisy and volatile due to inconsistent daily testing numbers. To generate a better realistic scenario in any developing country, the 5-day moving average of the daily confirmed cases has been used Ganyani et al., (2020). The daily growth factor of COVID-19 new infections has been calculated using the following expression with an aim to understand the progression of the novel coronavirus in any country.

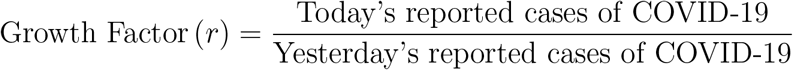

For any country, *r* > 1 depicts that the trend curve of new infections blows up exponentially, whereas *r* < 1 represents that the same curve decays to zero.

Daily incidence proportion (DIP) is the next epidemiological risk measure which has been used to estimate the probability of developing disease in any country Lai et al., (2020a) and Lai et al., (2020b). In an outbreak setting, from this estimation we can get a deeper understanding of disease progression as well as frequency of daily occurrence of new COVID-19 cases within a population as time progresses. It can also be referred as an attack rate or risk of developing any disease. As discussed earlier, a 5-day moving average of daily new infected cases has been used to calculate the DIP (per 1, 000, 000 per day) using the following expression:

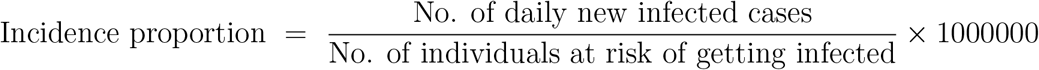

The denominator represents susceptible population in any country i.e. individuals who have the probability of getting infected with the disease. A high DIP depicts that the disease is transmitting into the population at an excessive rate. On the other hand, a low DIP indicates a minimal disease transmission in any community. In addition, disease incidence is positively correlated with mortality according to a recent study Lai et al., (2020b). Henceforth, to understand the severity of any disease outbreak, disease incidence proportion is an insightful epidemiological parameter.

Daily cumulative index (DCI) is used as one of the most significant indicators to understand the evolution dynamics of any epidemic. Recent studies Lai et al., (2020a) and Lai et al., (2020b) have enlightened the fact that this indicator could give robust understanding about the average daily number of new infected cases in a country. It has been observed in recent studies that DCI is significantly correlated with mortality compared to disease incidence Lai et al., (2020b). DCI can be defined as follows:

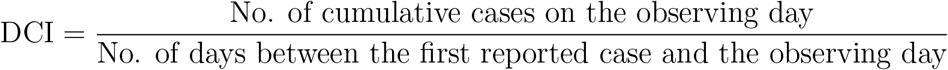

A continuous uptrend of DCI indicates that the average number of infected individuals is increasing sharply on a daily basis, whereas a constant declining trend of DCI depicts the fact that the mean value of daily infections is declining. A reduction in DCI can be achieved by imposing successful infection control measures. Henceforth, trend analysis of DCI plays a noticeable role in understanding the effectiveness of various disease control measures.

To quantify the transmissibility of an epidemic, effective reproduction number (*R*_*e*_) can be regraded as one of the most insightful epidemiological parameters. This significant metric represents the average number of secondary infections caused by an individual who has the capability to transmit any disease in an entirely susceptible population Nabi (2020). To date, sophisticated statistical methods have been employed to estimate the value of *R*_*e*_ precisely from the confirmed number of daily infected cases. Nevertheless, it is often really challenging to determine reliable estimates of *R*_*e*_ from available information. In our study, disease generation interval which refers to the duration between symptom onset of the primary case and the symptom onset of the secondary case has been used to estimate the effective reproduction number (*R*_*e*_) of COVID-19 Kohlberg & Abraham, (2020). In several recent studies Ganyani et al., (2020); Nishiura et al., (2020); Du et al., (2020), it has been found that the mean generation interval of COVID-19 is approximately 4 days. By using the generation interval of COVID-19, *R*_*e*_ can easily be calculated from daily new infections using the following expression Kohlberg & Abraham, (2020):

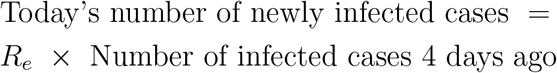

Consequently, *R*_*e*_ can be expressed as follows:

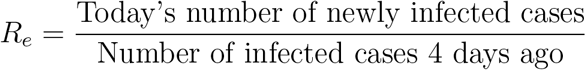

Generally, the true number of infected cases is relatively higher than the confirmed number of cases. Our estimation would not be affected for such discrepancies. For instance, if the actual number of cases is three times higher than the reported number of cases, then the multiplying factor will cancel out in the division. To smooth out the fluctuations in number of testing, a 5-day moving average of daily reported cases has been used in this study. A simple overview of disease burden can be gleaned using this basic approximation of the effective reproduction number in terms of two simple empirical parameters.

## 3. Numerical results and discussions

### 3.1. Bangladesh

On March 8, 2020, the very first case of COVID-19 was confirmed by the Institute of Epidemiology, Disease Control and Research (IEDCR) of Bangladesh Paul (2020, March 8). As the country witnessed the first death case due to the COVID-19, the government officials reacted swiftly and announced closures of all educational institutions from March 18 onwards. Afterwards, when there were 27 active COVID-19 cases out of 39 total cases, the government decided to enforce a countrywide lockdown to slow down the transmission of the virus with an exemption for the emergency services from March 26 Mithu & Ahmed (2020, March 24). This lockdown can be regarded as the most strict lockdown until the decision of reopening shops and malls maintaining health guidelines for limited time period came into force from May 10. This decision of strict lockdown was taken by the officials under immense pressure regarding the biggest Muslim festival (Eid-Ul-Fitr). It has been found in our study that the growth factor of new infections followed a downward trend and reached 0.3 before the lockdown could have been responsible for such decline (Fig. 1b). Disease incubation period (around 5 days Nabi (2020)) should be considered in this regard to determine the earliest effect of the imposed lockdown. Nevertheless, after five days of the lockdown a noticeable upswing of growth factor has been developed and hovered between 1.66 and 2 for one week (Fig. 1b). On the last day of this lockdown phase, the number of cumulative cases surpassed 14657. In addition, an average growth factor of 1.165 (95% confidence interval (CI): 1.08-1.25) in daily confirmed cases was conspicuous in this entire period (Table 1). In the second lockdown phase, it has been observed that the growth factor maintained a incessant trend (Mean: 1.05; SD: 0.05) even though the lockdown intensity has been lowered. Moreover, daily incidence proportion and daily cumulative index both got doubled in this period (Figs. 1d and 1e). After a 66-day countrywide lockdown period, government officials have been compelled to lift the lockdown with some health guidelines from June 1 considering the catastrophic economic recession Kamruzzaman (2020, May 28). The strict lockdown policy has been unsuccessful to force down the epidemic threshold below 1 (Fig. 1f). When a strict two-month long lockdown policy has failed to flatten the contagion curve, then the effectiveness of the most optimistic lockdown theories remains as an enigma.

**Table 1:** Growth factor of daily infected cases, daily incidence proportion, daily cumulative index and effective reproduction number in **Bangladesh** considering different lockdown phases from March 8 to June 12.

| Epidemiological Parameter | Before lockdown |  | Most strict lockdown<sup>a</sup> |  | Strict lockdown<sup>b</sup> |  | Unlockdown<sup>c</sup> |  |
| --- | --- | --- | --- | --- | --- | --- | --- | --- |
|  | Mean [95% CI] | SD | Mean [95% CI] | SD | Mean [95% CI] | SD | Mean [95% CI] | SD |
| Growth Factor (GF) | 1.31 [1, 1.62] | 0.5 | 1.165 [1.08, 1.25] | 0.305 | 1.05 [1.03, 1.076] | 0.054 | 1.03 [1.02, 1.04] | 0.02 |
| Daily Incidence Proportion (DIP) | 0.039 [0.023, 0.054] | 0.033 | 5.4 [4.1, 6.74] | 4.64 | 26.1 [23.2, 29.1] | 6.89 | 48.1 [45.4, 50.8] | 4.8 |
| Daily Cumulative Index (DCI) | 1.255 [0.9, 1.61] | 0.765 | 73.8 [52.3, 95.3] | 74.45 | 384.8 [343.1, 426.4] | 97.4 | 705.9 [657.9, 754] | 84.9 |
| Effective Reproduction Number | 2.78 [1.54, 4.01] | 2 | 2.244 [1.59, 2.89] | 2.25 | 1.22 [1.16, 1.27] | 0.13 | 1.15 [1.1, 1.2] | 0.09 |

<sup>a</sup> The most strict countrywide lockdown was effective from 26 March to 9 May.
<sup>b</sup> Strict lockdown was effective from May 10 to May 31.
<sup>c</sup> Lockdown has been lifted from June 1 onwards.

**Figure 1:**
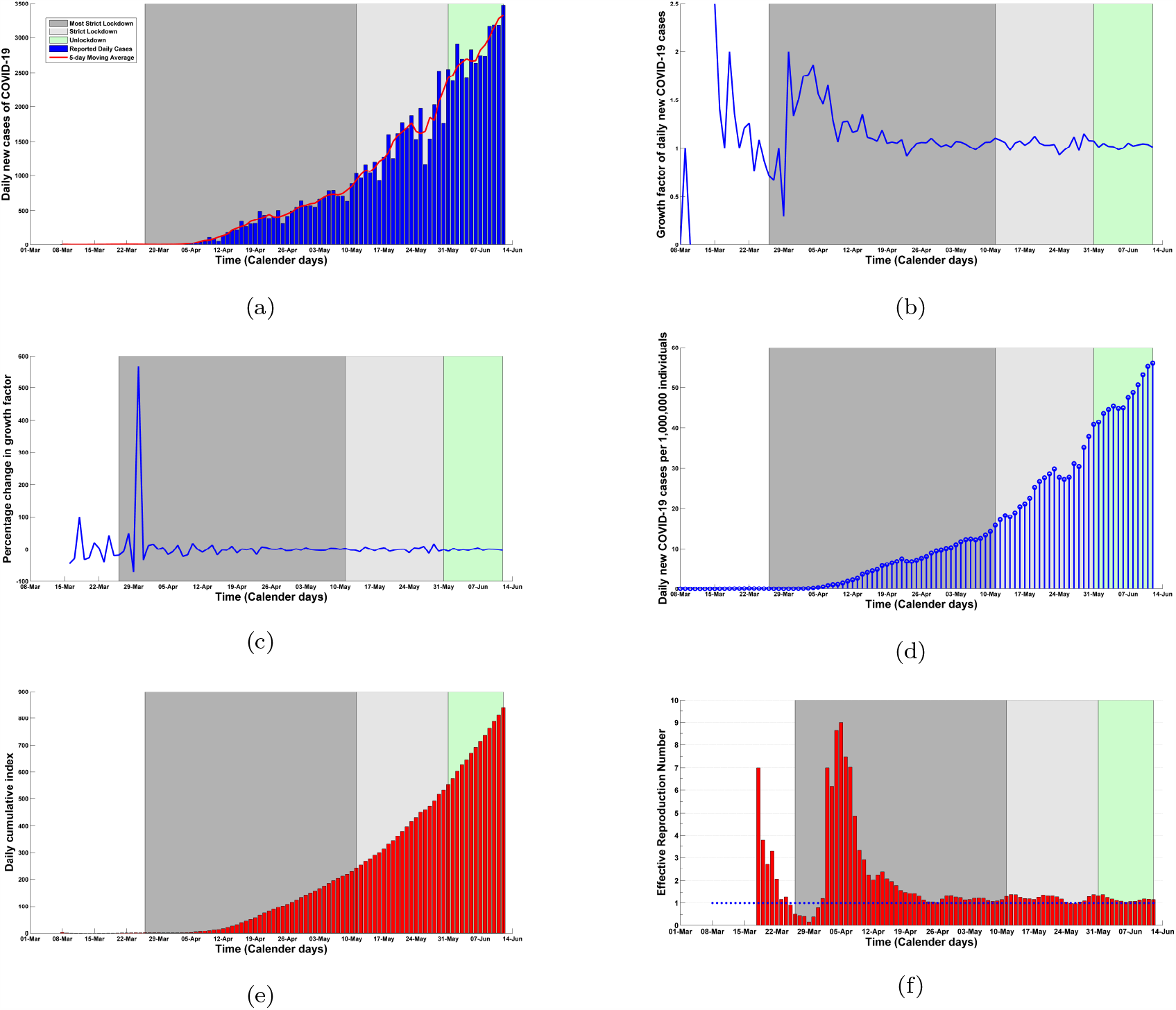
(a) Daily confirmed cases and 5-day moving average of daily cases, (b) growth factor, (c) percentage change in growth factor, (d) daily incidence proportion, (e) daily cumulative index and (f) effective reproduction number considering different lockdown phases in **Bangladesh** from March 8 to June 12. The intensities of different lockdown phases have been indicated in different colors: before lockdown (8/03/2020 – 25/03/2020); most strict lockdown (26/03/2020 – 09/05/2020); strict lockdown (10/05/2020 – 31/05/2020); unlockdown (1/06/2020 – 12/06/2020).

### 3.2. Brazil

Brazil is currently the second-worst COVID-19 affected countries in the world and has strong possibility to outnumber the tally of infected cases reported in the United States which is holding the leading position as of June 14, 2020. On February 25, a 61-year-old man who returned from Italy was reported as the first COVID-19 case in Brazil Siciliano et al., (2020). In response, Brazilian Health Ministry recommended at least 7-day isolation for both Brazilians and travellers arriving from outside. On March 23, officials of Rio de Janeiro imposed a strict lockdown in the state which is followed by Sao Paulo from the next day Mello & Gaier (2020, April 9). A downtrend in growth factor (GF) was evident before the enforcement of the strict sublockdown in Brazil (Fig. 2b). During the sublockdown phase, GF maintained a constant mean value of 1.08 (95% CI: 1.04-1.11) with standard deviation (SD) around 0.1 (Table 2). Brazil eased its lockdown from June 2 with a view to mitigating the deeper economic recession in the country Rivers & Gallón (2020, June 3). Despite the prolonged lockdown, the rate of growth of new infections was almost identical with negligible deviation from the mean value (Table 2). Galloping growths in both DIP and DCI were evident in different lockdown periods (Figs. 2d and 2e). Importantly, the effective reproduction number could not be brought under unity (Fig. 2f) in spite of having more than two months of lockdown (Table 2).

**Table 2:** Growth factor of daily infected cases, daily incidence proportion, daily cumulative index and effective reproduction number in **Brazil** considering different lockdown phases from February 25 to June 11.

| Epidemiological Parameter | Before lockdown |  | Strict sublockdown<sup>a</sup> |  | More strict sublockdown<sup>b</sup> |  | Easing of lockdown<sup>c</sup> |  |
| --- | --- | --- | --- | --- | --- | --- | --- | --- |
|  | Mean [95% CI] | SD | Mean [95% CI] | SD | Mean [95% CI] | SD | Mean [95% CI] | SD |
| Growth Factor (GF) | 1.44 [1.24, 1.65] | 0.49 | 1.08 [1.04, 1.11] | 0.1 | 1.06 [1.03, 1.09] | 0.08 | 1.03 [0.98, 1.088] | 0.09 |
| Daily Incidence Proportion (DIP) | 0.78 [0.27, 1.31] | 1.36 | 23 [18.28, 27.7] | 13.4 | 192.4 [159.6, 225.3] | 104.7 | 440.7 [413, 468.5] | 46.9 |
| Daily Cumulative Index (DCI) | 7.33 [2.9, 11.8] | 11.5 | 338.2 [259.8, 416.6] | 222.7 | 2477 [2070, 2880] | 1288 | 6240 [5860, 6630] | 649.1 |
| Effective Reproduction Number | 4.27 [2.96, 5.59] | 3.08 | 1.42 [1.29, 1.55] | 0.36 | 1.27 [1.21, 1.34] | 0.22 | 1.07 [0.99, 1.15] | 0.14 |

<sup>a</sup> First strict sublockdown was enforced on 23 March.
<sup>b</sup> More strict sublockdown was effective between 24 April and 1 June.
<sup>c</sup> Lockdown has been eased on 2 June.

**Figure 2:**
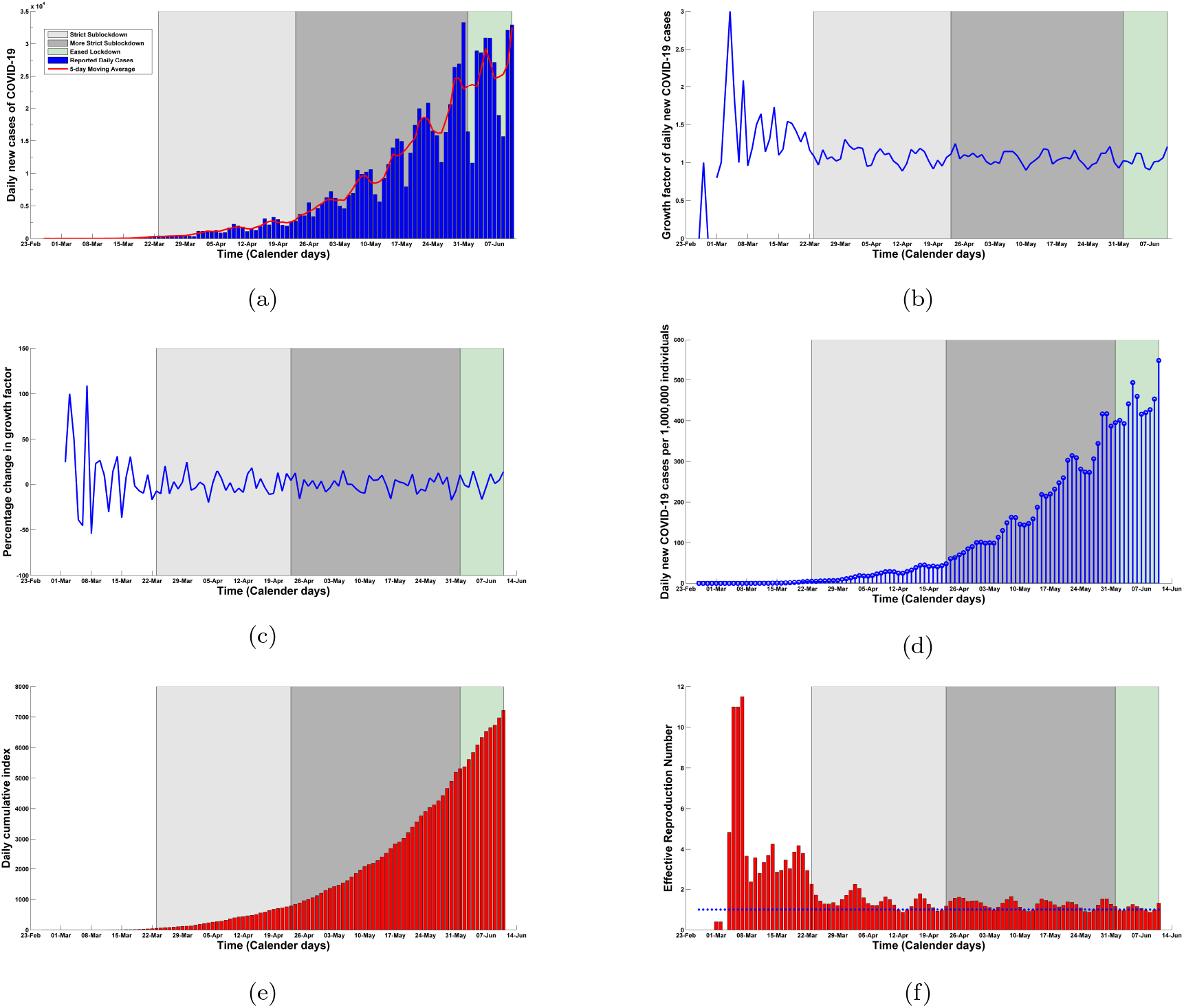
(a) Daily confirmed cases and 5-day moving average of daily cases, (b) growth factor, (c) percentage change in growth factor, (d) daily incidence proportion, (e) daily cumulative index and (f) effective reproduction number considering different lockdown phases in **Brazil** from February 25 to June 11. The intensities of different lockdown phases have been indicated in different colors: before lockdown (25/02/2020–22/03/2020); strict sublockdown (23/03/2020–23/04/2020); more strict lockdown (24/04/2020–1/06/2020); easing of lockdown (2/06/2020–11/06/2020).

### 3.3. Chile

The very first case of COVID-19 who had fresh travel history to Italy, was reported in Chile on March 3, 2020 Ministry of Health, Chile (2020). Afterwards, a faster growth rate of new infections (reached upto 2.3) was evident (Fig. 3b) and within 20 days the South American nation surpassed 632 confirmed COVID-19 cases Ministry of Health, Chile (2020). With an aim to stem the spread of coronavirus, a nationwide night-time 2200-0500 curfew was announced by the Chile’s government on March 22 Sherwood (2020, March 22). Fig. 3a illustrates that the daily reported infected cases and its 5-day moving average between March 3 and June 13. Before the nightly curfew, the mean growth factor of daily infections was estimated to be around 1.09 (95% CI: 1.05-1.13) (Table 3). Nevertheless, the number of daily infected cases continued to creep upwards incessantly and community transmission could not be checked in this curfew period. In consequence, Chile’s government ratcheted up the restrictive measures by introducing general mandatory quarantine for the citizens in an effort to force down the contagion rate. In this phase, growth factor maintained a continuous upward trend and reached a maximum value of 1.26 on May 1 (Fig. 3b). In addition, a sharp daily increase of about 22% in growth factor was noticeable in this curfew period (Fig. 3b). As of May 15, a complete shutdown of country’s capital Santiago was imposed as 80% of infected cases were identified in the capital Aljazeera (2020, May 14). Despite the strict curfew across the country, the daily cases per 1,000,000 individuals (daily incidence) soared up from 28.3 to 104.1 and the daily cumulative index shot up from 429 to 1580 within just 32 days (Figs. 3d and 3e). This continuous upswing in new infections raised questions about the efficacy of the most optimistic theories of lockdown. Importantly, Fig. 3f depicts that this strict confinement measures have failed to bring down the epidemic threshold below 1. The mean value of this insightful epidemiological parameter was 1.19 (95% CI: 1.14-1.26) in this above-mentioned period and according to our estimation this metric is going to maintain an upward trajectory in the upcoming days. As of June 13, Chileans are facing continuous spikes in daily cases and health care services are struggling to cope with this unforeseen crisis in spite of ongoing stringent curfew.

**Table 3:** Growth factor of daily infected cases, daily incidence proportion, daily cumulative index and effective reproduction number in **Chile** considering different lockdown phases from March 3 to June 13.

| Epidemiological Parameter | Before lockdown |  | Nationwide nightly curfew<sup>a</sup> |  | More strict curfew<sup>b</sup> |  | Shutdown of country’s capital<sup>c</sup> |  |
| --- | --- | --- | --- | --- | --- | --- | --- | --- |
|  | Mean [95% CI] | SD | Mean [95% CI] | SD | Mean [95% CI] | SD | Mean [95% CI] | SD |
| Growth Factor (GF) | 1.24 [1.02, 1.46] | 0.48 | 1.09 [1.05, 1.13] | 0.08 | 1.04 [1.02, 1.07] | 0.07 | 1.05 [1.03, 1.07] | 0.06 |
| Daily Incidence Proportion (DIP) | 0.41 [0.18, 0.64] | 0.49 | 4.35 [3.64, 5.05] | 1.48 | 11.87 [9.69, 14.06] | 6.5 | 67.67 [60.87, 74.47] | 19.9 |
| Daily Cumulative Index (DCI) | 6.44 [3.13, 9.76] | 7.16 | 80.2 [62.8, 97.5] | 36.5 | 248.7 [222.9, 274.5] | 76.7 | 962.9 [841.2, 1084] | 356.4 |
| Effective Reproduction Number | 2.64 [1.78, 3.5] | 1.86 | 1.49 [1.33, 1.64] | 0.33 | 1.2 [1.11, 1.29] | 0.27 | 1.19 [1.14, 1.26] | 0.18 |

<sup>a</sup> First nationwide night-time curfew was enforced on 22 March.
<sup>b</sup> More strict nationwide night-time curfew with mandatory quarantine restrictions was effective from 9 April and 14 May.
<sup>c</sup> Santiago which is the capital of Chile has been locked down on 15 May.

**Figure 3:**
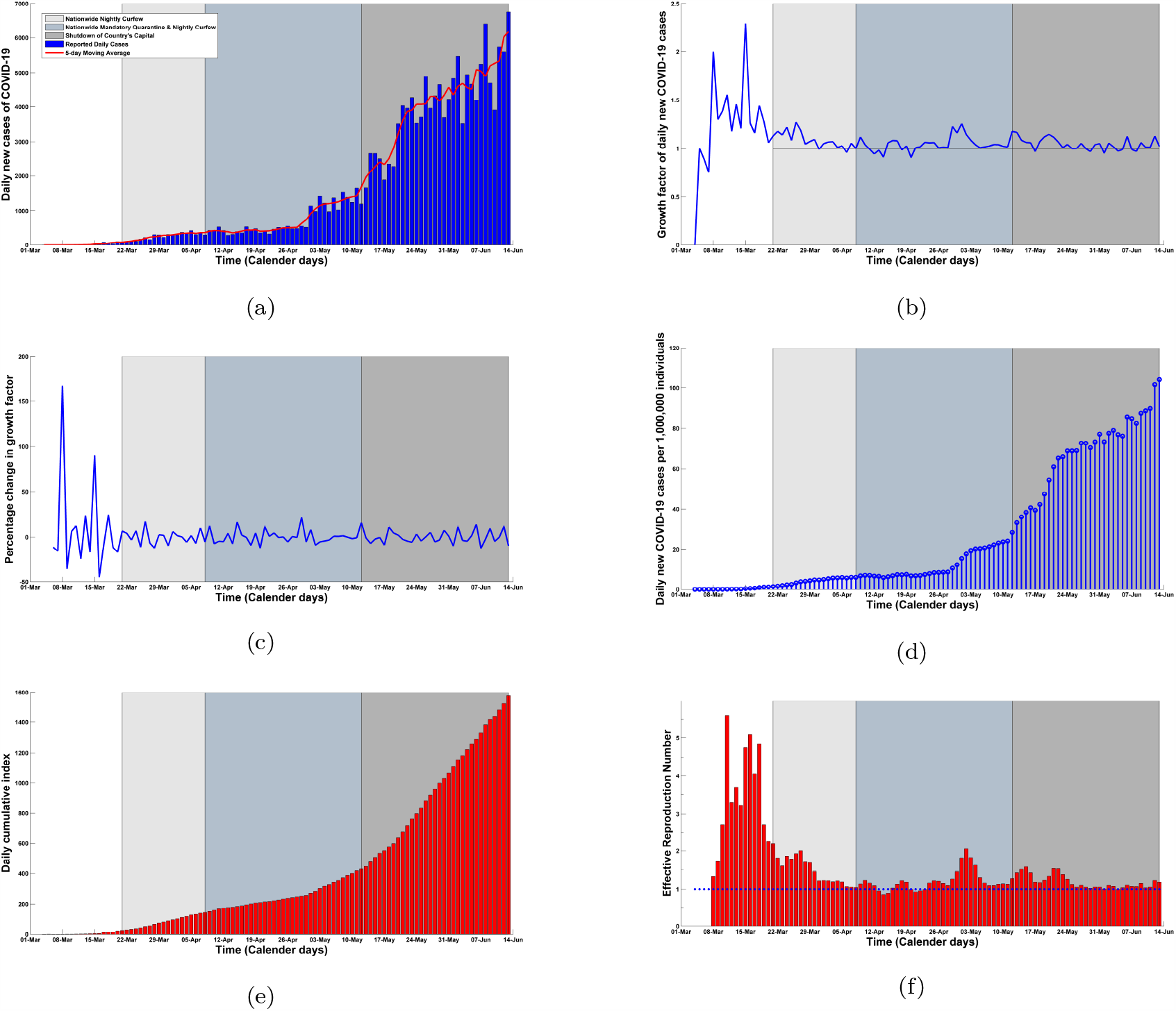
(a) Daily confirmed cases and 5-day moving average of daily cases, (b) growth factor, (c) percentage change in growth factor, (d) daily incidence proportion, (e) daily cumulative index and (f) effective reproduction number considering different lockdown phases in **Chile** from March 3 to June 13. The intensities of different lockdown phases have been indicated in different colors: before curfew (03/03/2020–21/03/2020); nationwide nightly curfew (22/03/2020–8/04/2020); more strict curfew (9/04/2020–14/05/2020); shutdown of country’s capital (15/05/2020–13/06/2020).

### 3.4. Pakistan

In Pakistan, a battle against the novel coronavirus has been started on February 26, with the identification of two COVID-19 positive cases who returned from Iran which was the worst-hit country at that time in Asian region Shahid (2020, February 27). With an aim to stanch the pandemic outbreak, a strict nationwide lockdown was forced by the officials of Pakistan in the middle of March Donaldson (2020, March 26). Growth factor had already begun to decline and reached 0.994 before the forced lockdown could have brought it under control (Fig. 4b). On March 31, government officials decided to lessen the intensity of the lockdown considering the economic recession and announced to open all kinds of transportation across the country Sajid & Latif (2020, March 24). During this partial lockdown period, daily incidence of COVID-19 ticked up from 1 to 28.3 and daily cumulative index also surpassed 376 following an upward trend (Figs. 4d and 4e). In this period, impoverished people started venturing out from their house in search of work as a significant number of people are dependent on daily wages. As the pandemic began to cripple Pakistan’s economy drastically, the government was compelled to take the decision of unlockdown with effect from May 9 Gulf Today (2020, May 9). Notably of all, in this unlockdown phase, it has been found in our analysis that the growth factor maintain an incessant trend with the mean value of around 1.04 (95% CI: 1.02-1.06, SD: 0.06) which is much lower than the average values of both the lockdown periods (Table 4). This enlightens that the highly debated unlockdown policy has not brought any significant change in the growth factor of new infections (Fig. 4c). Moreover, Fig. 4f illustrates that the effective reproduction number also plateaued during this unlockdown period (Mean: 1.19; 95% CI: 1.12-1.27) (Table 4). Social distancing measures and stringent mask wearing guidelines may have been the actual trigger for forcing down this crucial parameter.

**Table 4:** Growth factor of daily infected cases, daily incidence proportion, daily cumulative index and effective reproduction number in **Pakistan** considering different lockdown phases from February 26 to June 12.

| Epidemiological Parameter | Before lockdown |  | Forced lockdown<sup>a</sup> |  | Partial lockdown<sup>b</sup> |  | Unlockdown<sup>c</sup> |  |
| --- | --- | --- | --- | --- | --- | --- | --- | --- |
|  | Mean [95% CI] | SD | Mean [95% CI] | SD | Mean [95% CI] | SD | Mean [95% CI] | SD |
| Growth Factor (GF) | 1.52 [0.34, 2.7] | 2.48 | 1.76 [0.53, 3] | 2.5 | 1.07 [1.02, 1.12] | 0.16 | 1.04 [1.02, 1.06] | 0.06 |
| Daily Incidence Proportion (DIP) | 0.02 [0.015, 0.033] | 0.02 | 1.62 [1.34, 1.89] | 0.57 | 10.5 [8.46, 12.56] | 6.53 | 48.3 [40.2, 56.5] | 24.6 |
| Daily Cumulative Index (DCI) | 0.94 [0.75, 1.13] | 0.4 | 25.4 [18.1, 32.65] | 14.8 | 169.7 [141.96, 197.4] | 88.3 | 688.7 [613.7, 763.7] | 226.4 |
| Effective Reproduction Number | 2.68 [0.58, 4.77] | 4.4 | 5.63 [1.5, 9.76] | 8.4 | 1.34 [1.22, 1.46] | 0.39 | 1.19 [1.12, 1.27] | 0.22 |

<sup>a</sup> First lockdown was enacted from 15 March, 2020.
<sup>b</sup> Partial lockdown was effective from 31 March to 8 May.
<sup>c</sup> Lockdown has been lifted on 9 May.

**Figure 4:**
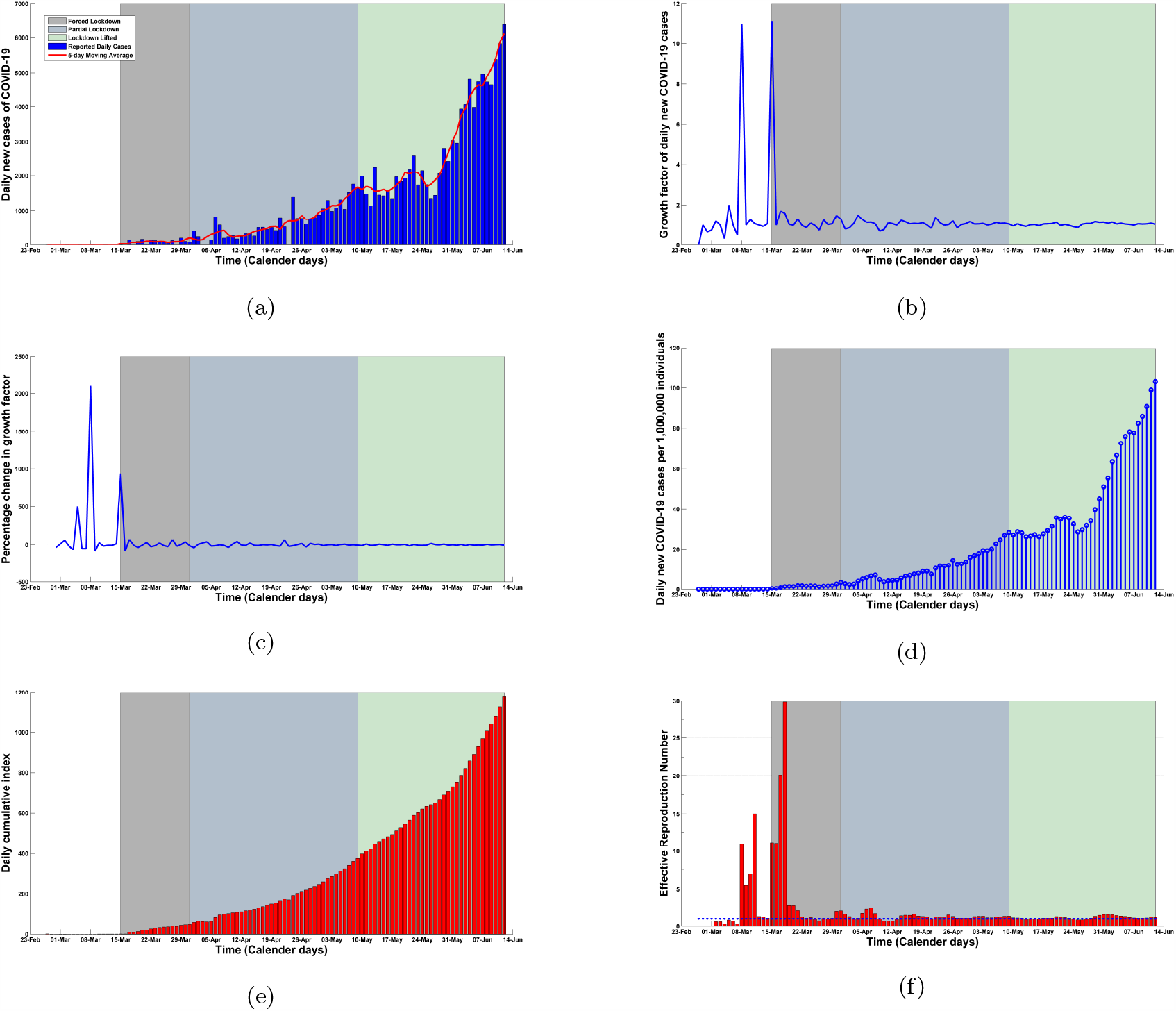
(a) Daily confirmed cases and 5-day moving average of daily cases, (b) growth factor, (c) percentage change in growth factor, (d) daily incidence proportion, (e) daily cumulative index and (f) effective reproduction number considering different lockdown phases in **Pakistan** from February 26 to June 12. The intensities of different lockdown phases have been indicated in different colors: before lockdown (26/02/2020–14/03/2020); forced lockdown (15/03/2020–30/03/2020); partial lockdown (31/03/2020–08/05/2020); unlockdown (09/05/2020–12/06/2020).

### 3.5. South Africa

In South Africa, the National institute for communicable diseases (NICD) first confirmed COVID-19 patient who returned back in South Africa from Italy on March 1, 2020 South African Ministry of Health (2020). To quell the progression of the outbreak, South Africa’s government launched a master plan of five-tier lockdown system which came into effect on March 26, 2020 Business Tech (2020, March 23). Before the imposition of the most strict (level-5) lockdown, the growth factor of new infections started to decline and reached its minimum value of 0.493 (Fig. 5b). Disease incubation period (around 5 days Nabi (2020)) should be considered in this regard to understand the earliest impact of the strict confinement measures. However, the growth rate began to soar and raised above 1 during this lockdown period (Table 5). After around one month, the entire country moved into a less strict lockdown (level-4) with stringent health protocols Voice of America (2020, March 14). Despite imposing harsh confinement measures, daily incidence continued to increase from 4.73 to 28.4 and daily cumulative index surged from 95.5 to 355 (Figs. 5d and 5e). As the economic pressure reached boiling point and several million people were struggling with hunger and managing their livelihoods, government have been compelled to ease the intensity of the lockdown from June 1 onwards Aljazeera (2020, May 25). In this new lockdown (level-3) period, epidemic threshold maintained an average value of 1.22 (95% CI: 1.11-1.32) with a small standard deviation of 0.17 (Table 5). In a nutshell, a two-month long lockdown has failed to bring the effective reproduction number under unity and bend the daily incidence curve as well (Figs. 5f and 5d). Despite the enforcement of stringent confinement policies, the outbreak situation is exacerbating as time progresses with new records in both infected and death cases.

**Table 5:** Growth factor of daily infected cases, daily incidence proportion, daily cumulative index and effective reproduction number in **South Africa** considering different lockdown phases from March 1 to June 10.

| Epidemiological Parameter | Before lockdown |  | Most strict lockdown<sup>a</sup> |  | More strict lockdown<sup>b</sup> |  | Strict lockdown<sup>c</sup> |  |
| --- | --- | --- | --- | --- | --- | --- | --- | --- |
|  | Mean [95% CI] | SD | Mean [95% CI] | SD | Mean [95% CI] | SD | Mean [95% CI] | SD |
| Growth Factor (GF) | 1.32 [1.11, 1.532] | 0.494 | 1.03 [0.97, 1.08] | 0.16 | 1.06 [1.04, 1.08] | 0.06 | 1.036 [0.988, 1.086] | 0.08 |
| Daily Incidence Proportion (DIP) | 0.6 [0.25, 0.96] | 0.84 | 2.21 [1.83, 2.596] | 1.15 | 13.9 [11.6, 16.23] | 6.58 | 37.22 [33.66, 40.78] | 5.74 |
| Daily Cumulative Index (DCI) | 8.29 [4.23, 12.35] | 9.49 | 62.23 [57.43, 67.04] | 14.5 | 201.4 [174.2, 228.6] | 77.2 | 455.2 [416.5, 493.9] | 62.4 |
| Effective Reproduction Number | 2.86 [2.08, 3.652] | 1.84 | 1.17 [1.05, 1.29] | 0.36 | 1.28 [1.23, 1.32] | 0.15 | 1.22 [1.11, 1.32] | 0.174 |

<sup>a</sup> The most strict national lockdown (Level 5) was imposed on 26 March.
<sup>b</sup> Stricter lockdown (Level 4) was effective from 1 May to 31 May.
<sup>c</sup> National restrictions has been eased (Level 3) on 1 June.

**Figure 5:**
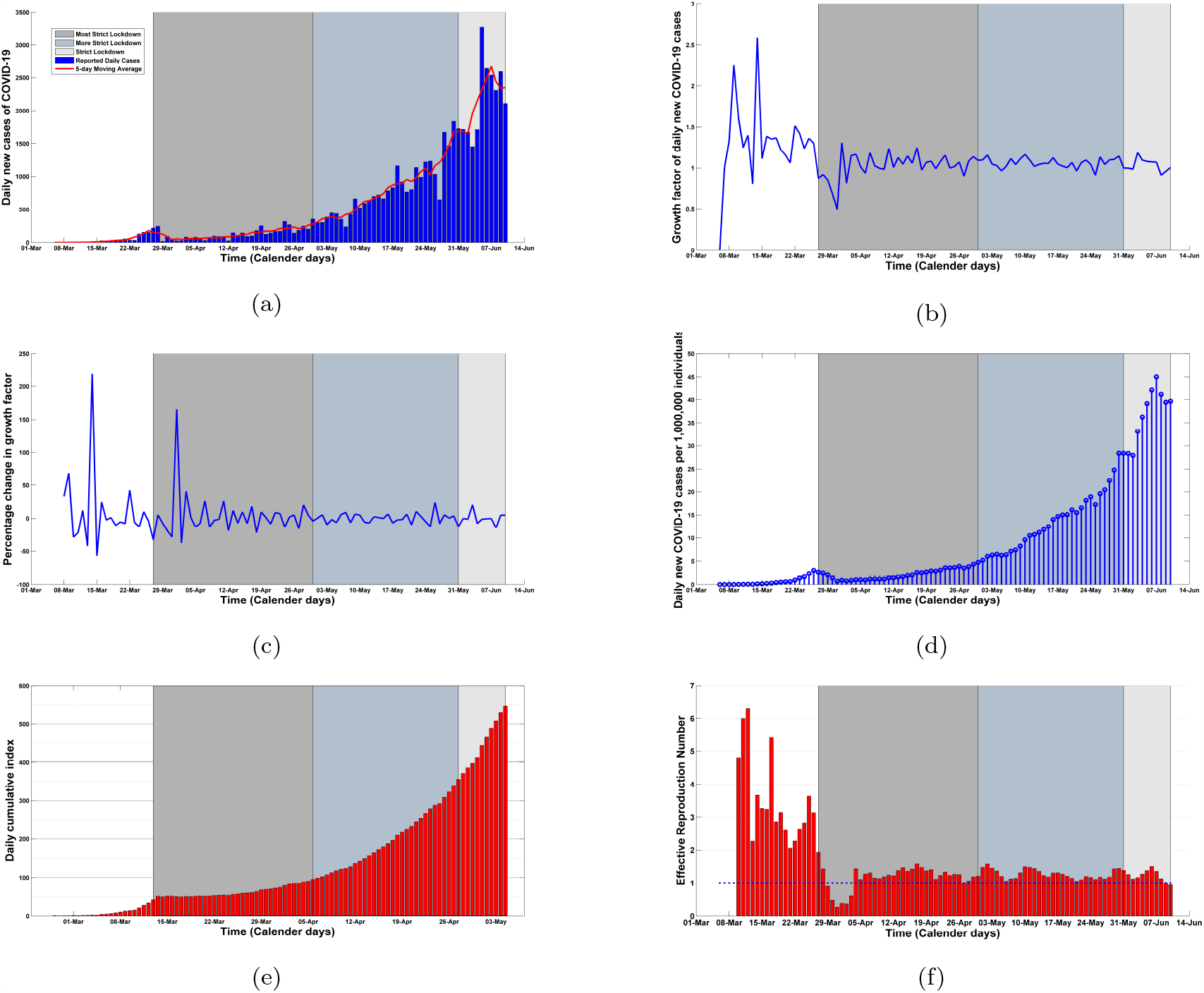
(a) Daily confirmed cases and 5-day moving average of daily cases, (b) growth factor, (c) percentage change in growth factor, (d) daily incidence proportion, (e) daily cumulative index and (f) effective reproduction number considering different lockdown phases in **South Africa** from March 1 to June 10. The intensities of different lockdown phases have been indicated in different colors: before lockdown (1/03/2020–25/03/2020); strictest lockdown (26/03/2020–30/04/2020); stricter lockdown (1/05/2020–31/05/2020); strict lockdown (1/06/2020–10/06/2020).

## 4. Concluding remarks

As the panic of the COVID-19 pandemic started unfolding to different parts of the world, developing countries hurriedly launched strict lockdown policies considering lockdown of Wuhan as a perfect model with an aim to quell the outbreak of the novel coronavirus. Unfortunately, such draconian measures could not be implemented successfully in developing countries compared to China because of unsuccessful isolation of country’s capital from other cities and strict confinement measures have also been unsuccessful owing to irresponsible behavior of the citizens in maintaining social distancing guidelines. It is also worth mentioning that the national economies of these developing countries comprehensively rely on their respective capital cities. For this reason, governments were unable to cut off the connectivity between country’s capital and other cities. After analyzing various trends of different epidemiological parameters in different lockdown phases, it has been unearthed that the impact of strict country-wide lockdowns for prolonged period is paper-thin in developing countries which might ignite a firestorm of controversy in the research community. Nevertheless, a rigorous real-time data analysis of five developing countries reveals the fact that those imposed lockdowns were not as effective to curb the spread of the virus as those were expected to be. On the other hand, inhabitants of these countries are facing unprecedented crises due to immense disruption of their respective national economies. As prolonged lockdown cannot be a pragmatic solution for developing countries due to its catastrophic socio-economic consequences, most of the respective governments have been compelled to ease the lockdown measures eventually. Our study suggests that a refinement of policy-making is indispensable for developing countries to ensure a perfect balance between saving lives and confirming livelihoods.

## Data Availability

Available online

https://ourworldindata.org/coronavirus-source-data

## References

Ahmed, Z., Coronavirus: Economy down, poverty up in Bangladesh, Deutsche Welle, June 10, 2020, Available Form: https://www.dw.com/en/coronavirus-economy-down-poverty-up-in-bangladesh/a-53759686.

Donaldson, T., Pakistan lockdown idles factories—where orders had shriveled up, Sourcing Journal, March 26, 2020. Available from: https://sourcingjournal.com/topics/sourcing/pakistan-coronavirus-lockdowngarment-factories-closed-synergies-worldwide-levis-202444.

Du, Z., Xu, X., Wu, Y., Wang, L., Cowling, B. J., & Meyers, L. (2020). Serial Interval of COVID-19 among Publicly Reported Confirmed Cases. Emerging Infectious Diseases, 26(6), 1341–1343.

Ganyani, T., Kremer, C., Chen, D., Torneri, A., Christel, F., Wallinga, J., Niel, H. (2020). Estimating the generation interval for coronavirus disease (COVID-19) based on symptom onset data, Euro Surveillance, 25(17).

Gollier, C. (2020). If the objective is herd immunity, on whom should it be built? EconPol Europe Policy Brief. Available Form: https://www.econpol.eu/publications/policy_brief_29.

Guilherme, D., Bruno, S., Bruno, B. F., Cleyton, M. S., Graciela, A. (2020). The impact of COVID-19 partial lockdown on the air quality of the city of Rio de Janeiro, Brazil, Science of The Total Environment, 729, 139085.

ICQI (2020), List of developing countries, International Congress of Qualitative Inquiry (Accessed on June 10, 2020), Available Form: https://icqi.org/developing-countries-list.

Kamruzzaman, M., Bangladesh to partly ease lockdown amid virus concerns, May 28, 2020, Available from: https://www.aa.com.tr/en/asia-pacific/bangladesh-to-partly-ease-lockdown-amid-virus-concerns/1855714.

Kohlberg, E., and Abraham, N. (2020). Demystifying the Math of the Coronavirus. emphHarvard Business School Working Paper, No. 20–112.

Lai, C., Wang, C., Wang, Y., Hsueh, S., Ko, W., & Hsueh, P. (2020). Global epidemiology of coronavirus disease 2019 (COVID-19): disease incidence, daily cumulative index, mortality, and their association with country healthcare resources and economic status. International Journal of Antimicrobial Agents, 55(4), 105946.

Lai, C., Wang, C., Wang, Y., & Hsueh, P. (2020). Global coronavirus disease 2019: What has daily cumulative index taught us? International Journal of Antimicrobial Agents, 55(6), 106001.

Lau, H., Khosrawipour, V., Kocbach, P., Mikolajczyk, A., Schubert, J., Bania, J., Khosrawipour, T. (2020). The positive impact of lockdown in Wuhan on containing the COVID-19 outbreak in China, Journal of Travel Medicine, 27(3).

Mahmood, M., COVID-19 and the developing world, The Financial Express, May 30, 2020, Available Form: https://thefinancialexpress.com.bd/views/reviews/covid-19-and-the-developing-world-1590852422.

Mello, G., & Gaier, R. V., “Brazil lockdowns, attacked by Bolsonaro, begin to slip”, April 9, 2020, Reuters, Available from: https://www.reuters.com/article/us-health-coronavirus-brazil-lockdown/brazil-lockdowns-attacked-by-bolsonaro-begin-to-slip-idUSKCN21R30B.

Meunier, T. A. J. (2020). Full lockdown policies in Western Europe countries have no evident impacts on the COVID-19 epidemic, MedRxiv, Available Form: https://doi.org/10.1101/2020.04.24.20078717.

Ministerio de Salud de Chile (Spanish), “Informe Epidemiológico COVID-19” (Accessed on June 10, 2020). Available from: https://www.minsal.cl.

Mithu, A. I., & Ahmed, M. F., “Bangladesh prepares for lockdown”, March 24, 2020, Available from:https://tbsnews.net/coronavirus-chronicle/coronavirus-bangladesh/bangladesh-prepares-lockdown-60595.

Nabi, K. N. (2020). Forecasting COVID-19 pandemic: A data-driven analysis. Chaos, Solitons & Fractals. Available from: https://authors.elsevier.com/tracking/article/details.doaid=110046&jid=CHAOS&surname=Khondoker.

Nishiura, H., Linton, N. M., Akhmetzhanov, A. R. (2020). Serial interval of novel coronavirus (COVID-19) infections. International Journal of Infectious Diseases, 93, 284–286.

Our World in Data (2020),”Coronavirus Source Data”, Available Form: https://ourworldindata.org/coronavirus-source-data.

Pakistan lifts lockdown amid jump in coronavirus cases, Gulf Today, May 9, 2020. Available from: https://www.gulftoday.ae/news/2020/05/09/pakistan-lifts-lockdown-amid-jump-in-coronavirus-cases.

Paul, R., Bangladesh confirms its first three cases of coronavirus, Reuters, March 8, 2020. Available from: https://www.reuters.com/article/us-health-coronavirus-bangladesh-idUSKBN20V0FS.

Ramaphosa: South Africa coronavirus lockdown to ease from June 1, Aljazeera, May 25, 2020. Available from: https://www.aljazeera.com/news/2020/05/ramaphosa-south-africa-coronavirus-lockdown-ease-june-1-200525070634938.html.

Ramaphosa announces 21 day coronavirus lockdown for South Africa, Business Tech, March 23, 2020. Available from: https://businesstech.co.za/news/government/383927/ramaphosa-announces-21-day-coronavirus-lockdown-for-south-africa.

Rivers, M., & Gallón, N., Mexico and parts of Brazil reopen after lockdown-despite surging coronavirus cases, CNN June 3, 2020. Available from: https://edition.cnn.com/2020/06/02/americas/latin-america-coronavirus-reopening-intl/index.html.

Sajid, I. & Latif, A., Pakistan stays under lockdown amid coronavirus outbreak, Anadolu Agency, March 24, 2020. Available from: https://www.aa.com.tr/en/asia-pacific/pakistan-stays-under-lockdown-amid-coronavirus-outbreak/1777394.

Shahid, A., Two coronavirus cases confirmed in Pakistan, Pakistan Today, February 27, 2020. Available from: https://www.pakistantoday.com.pk/2020/02/26/sindh-health-two-coronavirus-cases-confirmed-in-pakistan-confirms-first-coronavirus-case-in-karachi.

Sherwood, D., Chile announces nationwide nightly curfew, coronavirus cases hit 632, National post, March 22, 2020. Available from: https://nationalpost.com/pmn/health-pmn/chile-announces-nationwide-nightly-curfew-coronavirus-cases-hit-632.

South African Ministry of Health, “Health updates on Coronavirus” (Accessed on June 10, 2020). Available from: https://www.gov.za/speeches/health-updates-coronavirus-10-mar-2020-0000.

South Africa set to move toward easing lockdown restrictions, Voice of America News, March 14, 2020. Available from: https://www.voanews.com/covid-19-pandemic/south-africa-set-move-toward-easing-lockdown-restrictions.

Tang, Y., and Wang, S. (2020). Mathematic modeling of COVID-19 in the United States. emphEmerging Microbes & Infections, 9(1), 827–829.

Total lockdown for Chile capital after surge in virus cases, Aljazeera, May 14, 2020. Available from: https://www.aljazeera.com/news/2020/05/total-lockdown-chile-capital-surge-virus-cases-200513163148849.

UN COVID-19 response. Available from: https://www.un.org/sustainabledevelopment/poverty.

Siciliano, B., Dantas, G., Silva, C. M., Arbilla, G. (2020). Increased ozone levels during the COVID-19 lockdown: Analysis for the city of Rio de Janeiro, Brazil. emphScience of the Total Environment, 737, 139765.

